# “It’s not about the money, money” - Well, actually it is. Divergent views on drivers of early phase clinical trial participation among ethnically diverse potential trial participants in the United Kingdom: A Mixed Methods Study

**DOI:** 10.1101/2024.04.04.24305355

**Authors:** Pilar Artiach Hortelano, Neil Morton, Paul Wicks, Michael Young, Rebecca Burdell, Duncan Richards

## Abstract

**Background:** Novel therapeutics should always be tested in a sample representative of the population in need of treatment. Initial efforts of drug development take place in early phase trials (phase-I and -II), setting the direction for late-stage studies (phase-III and -IV). However, study samples in early phase trials typically fail to recruit Black, Asian and minority ethnic groups, which might produce results which don’t generalise to a broader population in later trials, and ultimately, clinical practice. Focusing on early phase clinical trials the present study (1) explored the barriers and incentives that determine participation of ethnic minorities in clinical research, and (2) proposes strategies that mitigate such barriers.

**Methods:** A systematic literature review explored barriers affecting participation rates from individuals from diverse ethnic backgrounds. An exploratory phase involved two online surveys (researchers and general population) and focus groups (general population) analysed using thematic analysis.

**Results:** The systematic review found little published evidence, with most studies undertaken in the USA and focused on specific clinical areas. The exploratory phase showed a discordance between researchers’ and general public’s perspectives on both drivers and barriers to early phase trial participation.

**Discussion:** These findings were synthesised into a *Clinical Trials Participatory Framework*, which contextualises reasons for reduced trial participation, while providing mechanisms/strategies to increase uptake among minority ethnic participants. This may guide researchers when implementing strategies to aid under-representation in their samples. Further research should evaluate the framework by actively implementing, testing, and iterating upon the strategies.

## Background

Novel therapeutics should be tested in samples that represent the population that will ultimately receive the treatment if licensed (1). However, for a long time there has been an underrepresentation of Black, Asian and other minority ethnic groups in clinical trials (2-4). This increases the risk of safety issues disproportionately affecting certain ethnic groups not being identified during development (5-9). Accordingly, there is a need to broaden participation in clinical research, specifically in early phase studies (Phase-I and -II).

Early phase, so-called ‘learning’ clinical trials, aim to provide an initial evaluation of safety and efficacy, setting the direction for future late stage ‘confirming’ studies. These studies, which might involve patients and healthy volunteers, must be able to provide robust conclusions that can guide and accurately determine progression into phase-III trials (10). Failure to adequately represent the target population in these studies exposes future patients to potential increased risk of both adverse effects and treatment failure, ultimately limiting utility in clinical practice.

To date, efforts to increase participation from diverse ethnic backgrounds has focussed on later-stage large scale research (Phase-III and -IV), has been based in the United States of America (USA) (11-13) and mostly focused on specific clinical areas (i.e., cancer (11) or asthma (14)). Recent early phase evidence has proposed reasons such as socio-economic status, lack of trust, previous negative experiences with the healthcare system and lack of appropriate communication as potential barriers impacting participation of ethnic minority groups (2). Given the paucity of evidence, further action is necessary to better understand these barriers and what can be done to mitigate them.

The present project, with a specific focus in early phase clinical trials (phase-I and -II), aimed to (i) explore the barriers precluding individuals from diverse ethnic backgrounds from partaking in clinical research, and (ii) find suitable strategies that would enhance the participation rate from these groups. There were two principal components: (i) a systematic review of the literature (ii) an exploratory phase involving two online e-surveys and focus groups. The e-surveys consisted of: (a) a *Researchers Survey* which explored researcher awareness of the lack of diversity in clinical research, and strategies that are being adopted to aid this; and (b) a *General Population Survey* to understand what incentives and barriers impact clinical trial participation in different ethnic groups. Finally, focus groups with members of diverse ethnic groups explored what strategies might make clinical research more accessible.

## Methods

### Systematic Review

The selection criteria specified barriers to diversity of participants in trials of new therapeutics compared to placebo/standard of care, in the early phase and within the last 20 years (i.e., 2002-2022). The time limit of 20 years was considered appropriate given the lack of focus on these issues prior to this. Specifically, barriers rather than enablers/solutions were looked for, and in interventional trials. A variety of phrases were used, seeking to encompass both Phase-I and Phase-IIa.

The search included word-directed literature search of primarily Pubmed, Medline, Cochrane and Google Search databases, using Boolean operators, for example: “(((barriers to diversity) AND (clinical trial)) AND (early phase)” and “((barriers to diversity) AND (clinical trial)) AND (ethnicity)”. Studies were selected for inclusion based on a review of keywords and the abstract confirming fit. Narrative synthesis was used for analysis of these searches.

### e-Surveys

#### Preparation and dissemination of e-Surveys

The *Researcher’s Survey* consisted of a 7-question survey (Supplementary Appendix 1) including structured (multiple choice) and unstructured (open answer) responses from researchers and principal investigators from different fields involved in participant-facing research. The survey collected background information about researchers (i.e., role they hold within the scientific community and phase of clinical trials they are involved in) and explored which demographic information was routinely collected and their perception of participant barriers to trial participation. The survey also asked what training or advice researchers had received on recruitment methods, the previous strategies employed to enhance diversity, and how participant language barriers (if present) were accommodated.

The *General Population Survey* consisted of a 14-question survey (Supplementary Appendix 2) for potential trial participants in the general population. Informed consent was obtained from participants prior to commencing the survey and contact details of a member of the research team were provided. Firstly, questions recorded demographic information about the participants (i.e., age, gender, ethnic background, level of education and employment). In the first part of the survey, a definition in lay language of a clinical trial was provided and later questions explored previous participation in clinical trials as well as willingness to take part in the future. Sequentially, incentives and barriers for taking part in clinical trials were explored. One item explored how much money participants would expect to be paid for taking part in a clinical trial. In the second part of the survey, a lay definition of early phase clinical trials was presented and, various items explored prior participation, willingness to take part in one in the future, and incentives and barriers to participation.

Surveys were designed with the input of the Enhancing Diversity in Clinical Trials (EDICT) advisory committee and fielded via Jotform (https://www.jotform.com). The *Researcher’s Survey* was disseminated through email to different clinical trial units across the UK, and research centres focused on participant-facing research. The *General Population Survey* was disseminated online in collaboration with Prolific (https://www.prolific.co) to ensure a diverse population was reached. Informed consent was obtained from participants prior to commencing the survey and contact details of a member of the research team were provided. The surveys were anonymized with no personal information collected. Participants were able to finalise the survey at any point. Data were collected through the online platform and exported to a csv file which only members of the research could access.

#### Data analysis

An exploratory analysis was carried with the data from both surveys. Data was initially collated, categorised and plotted for visual inspection, followed by a descriptive analysis.

### Focus groups

#### Procedure

A targeted social media advertising campaign was run by Lindus Health to find willing participants for the focus groups. A short Jotform survey (https://www.jotform.com) was linked to the social media campaign to assess willingness to partake in a focus group. Further background information was gathered from interested participants (i.e., age, gender, ethnicity, education level, current employment, past trial experience and contact details for organisation purposes). Potential participants gave informed consent and authorisation for the meetings to be recorded before participation. Personal data was stored securely such that it was only accessible to members of the research team.

Interested participants were asked for their preferred date and time for the focus group and then assigned by convenience to one. In total, 17 participants were recruited and divided into five groups. The duration of the focus group was 60 minutes, and participants were reimbursed with £20 for their participation. Focus groups were led by one moderator (male or female) and an observer was present.

In preparation for the focus groups, participants were given a document where general instructions and dynamics for the meeting were provided (Supplementary Appendix 3). This included a glossary with a lay definition of terms that were going to be used throughout the meeting (i.e., clinical trial, placebo, etc.,). At the start of the meeting, the moderator introduced the dynamics of the focus group, and both the moderator and observer introduced themselves. The moderator then guided and stimulated the debate amongst participants to obtain their views on clinical trials, with a specific focus on early phase clinical trials, incentives and barriers that would determine their participation in one, and what strategies could be put in place to enhance the incentives and overcome the barriers to boost participation. Video and audio were recorded throughout the meeting.

#### Data collection and analysis

Data obtained from the focus groups was analysed using Kiger and Varpio’s (15) guide for thematic analysis. The researcher was familiarised with the focus groups recordings, and transcribed the recordings assigning a code to each participant to preserve anonymity. Following transcription, each assessment was verified with the observer to ensure comprehensiveness of the data and transcripts were subjected to initial coding by the researcher, preserving participant’s language and gerunds. Recurring initial codes were subsumed into themes reflecting factors that may potentiate trial participation, taking into consideration existing barriers and strategies to overcome these.

### Ethics

It was determined by the project steering committee that an ethics committee application was not needed for the online surveys nor focus groups as these fall under the category of “market research”. This was informed using the HRA Research Decision Tool (http://www.hra-decisiontools.org.uk/research/).

## Results

### Systematic review

The literature searches yielded over 100 results with mention of barriers to diversity based on clinical trial data, but less than 10 focused on barriers to diversity specifically in interventional trials. When identifiers phrases relating to early phase/Phase-I/Phase-IIa were included, there were no reviews focussing specifically on these issues. There were just 4 studies focussing on barriers to participation of ethnic minorities (n=4) which are discussed below.

Trials within the 20-year limit highlighted studies based in the USA beginning to focus on barriers to participation of ethnic minorities (16). These included categories of barriers and highlighted time pressure for participants as the biggest barrier. Many of the remaining search results were studies focussing on a particular clinical area, and therefore although early phase trials were mentioned, barriers identified were not all applicable for consideration of barriers for healthy volunteers. Nevertheless, useful, and consistent themes were identified. Clark and colleagues (17) focussed on four critical categories of barriers to participation but did focus specifically in African/Caribbean/Hispanic populations in the USA rather than generalising barriers to diversity participation in other populations. These barriers were categorised as *mistrust*, *lack of comfort with process*, *time/resource constraints*, and *lack of awareness*. The most detailed analyses of barriers came from large cancer studies (18, 19). These grouped factors into individual/patient-based, systematic (such as relating to cost and the trial itself), and interpersonal (i.e., doctor-patient relationship) factors. These large studies used input from different stakeholders including patients from minorities, referring physicians, and investigators. Although studies focussing on barriers to participation were plentiful, those relating to early phase trials were very limited.

In summary there was limited existing literature focussing on the specific barriers to diversity in early phase clinical trials, however the identified themes may be used to inform further work in the field.

### Surveys

#### Researchers Survey

##### Participants

A total of 53 researchers responded to the survey. Researcher’s background can be found in Table 1. 30.19% of responders reported their work to pertain to early phase clinical trials (i.e., phase-I and -II); and 39.62% to late phase clinical trials (i.e., phase-III and -IV). The remainder of the sample (30.19%) did not identify as working in a specific phase within clinical trials or did other types of human-facing research.

**Table 1.**
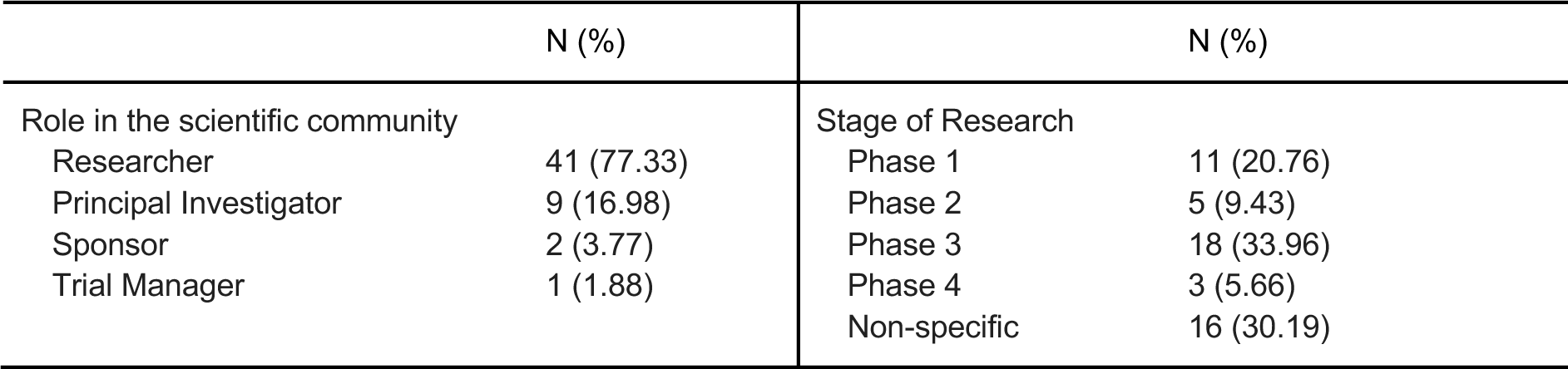
Researcher’s background.

##### Common demographic information collected from participants

Across the sample, age and gender were most collected in trials (94.3% and 92.5% of researchers, respectively). This was followed by 52.8% of the sample asking for ethnic background, and 45.3% for years of education. Finally, 7.6% requested civil status, 5.7% for country of origin and 5.7% for “other”. No respondents are asking for religion. Differences in demographic information acquired between early phase and late phase researchers can be observed in Figure 1a. Interestingly, in early phase clinical trials 68.75% of researchers ask for ethnic background, compared to 57.1% in late phase clinical trials. Similarly, a higher proportion of researchers in early phases ask for country of origin (12.5%), compared to those in later phases (4.8%).

**Fig 1.**
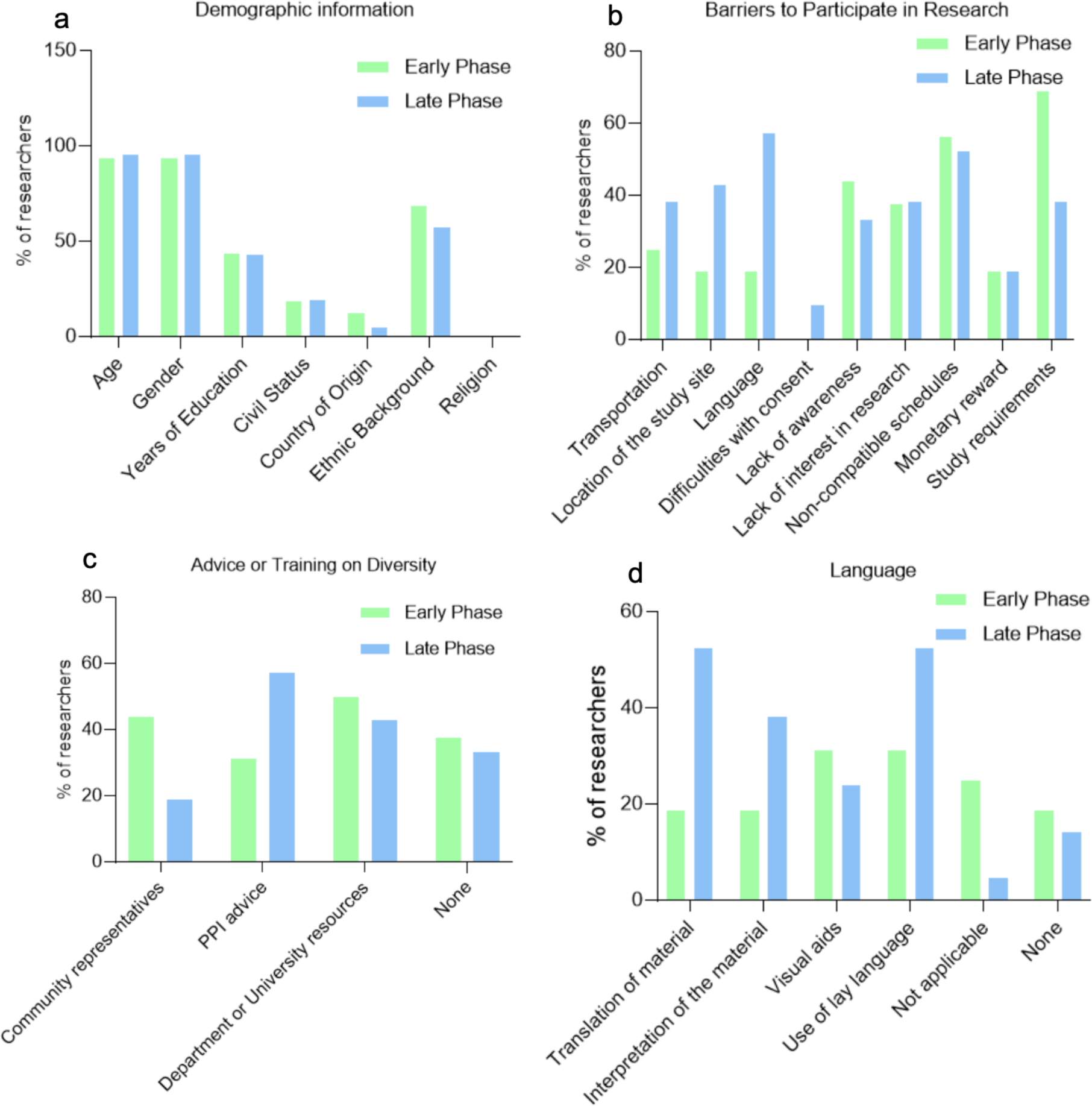
Difference in responses between early phase and late phase researchers. (a) Demographic information collected from respondents pertaining to early and late phase clinical trials. (b) Barriers to participation identified by early and late phase researchers. (c) Advice or training received on recruitment methods to improve diversity according to early or late phase researchers. (d) Strategies to tackle language barriers according to early and late phase researchers.

##### Barriers for taking part in research

Survey responders were asked to select the three most common barriers they felt precluded participants from taking part in their research from a predefined list. Across the sample, the three most common barriers for partaking in research were (1) *complicated study requirements* (49%), (2) *non-compatible schedules* (45.3%), and (3) *difficulties with consent/Lack of awareness/Lack of interest in science* (41.5%). The least mentioned barrier for researchers was *poor or lack of monetary reward* (15.1%). Differences in responses between early and late phase researchers can be seen in Figure 1b.

##### Advice or training received on recruitment methods to improve diversity

Overall, 36% of our researcher sample did not seek advice on recruitment methods to improve the diversity of their sample. Among those who did, advice is mostly obtained from their department or university, or Patient and Public Involvement (PPI) groups (43.4%). Differences in early and late phase can be found in Figure 1c.

##### Strategies employed to encourage more participants from diverse backgrounds

Overall, 47.2% did not implement any strategies to encourage a more diverse sample. This percentage was the highest in early phase research (62.5%), and lowest in later phases (33.3%). The most common strategies were (1) *strategic location of study site/advertisement to target different ethnic groups*, (2) *translation of material* and (3) *use of social media*. In early phase research, one researcher (6.25%) reported using *community engagement strategies* and another one (6.25%) *advice from PPI groups*.

##### Overcoming language barriers

Overall, 24.53% of responders reported language not to be a barrier in their research and 5.67% of researchers reported knowledge of English as a requisite for participation. Among those who reported employing strategies for language as a barrier for participation, *translation of material* was the most widely used strategy (43.4%), followed by *use of lay language* (37.3%). Differences in strategies employed between researchers in early and late phase clinical trials can be found in Figure 1d.

#### General Population Survey

##### Participants

A total of 1049 individuals from the general population answered the survey. Their demographic information can be found in Table 2.

**Table 2.**
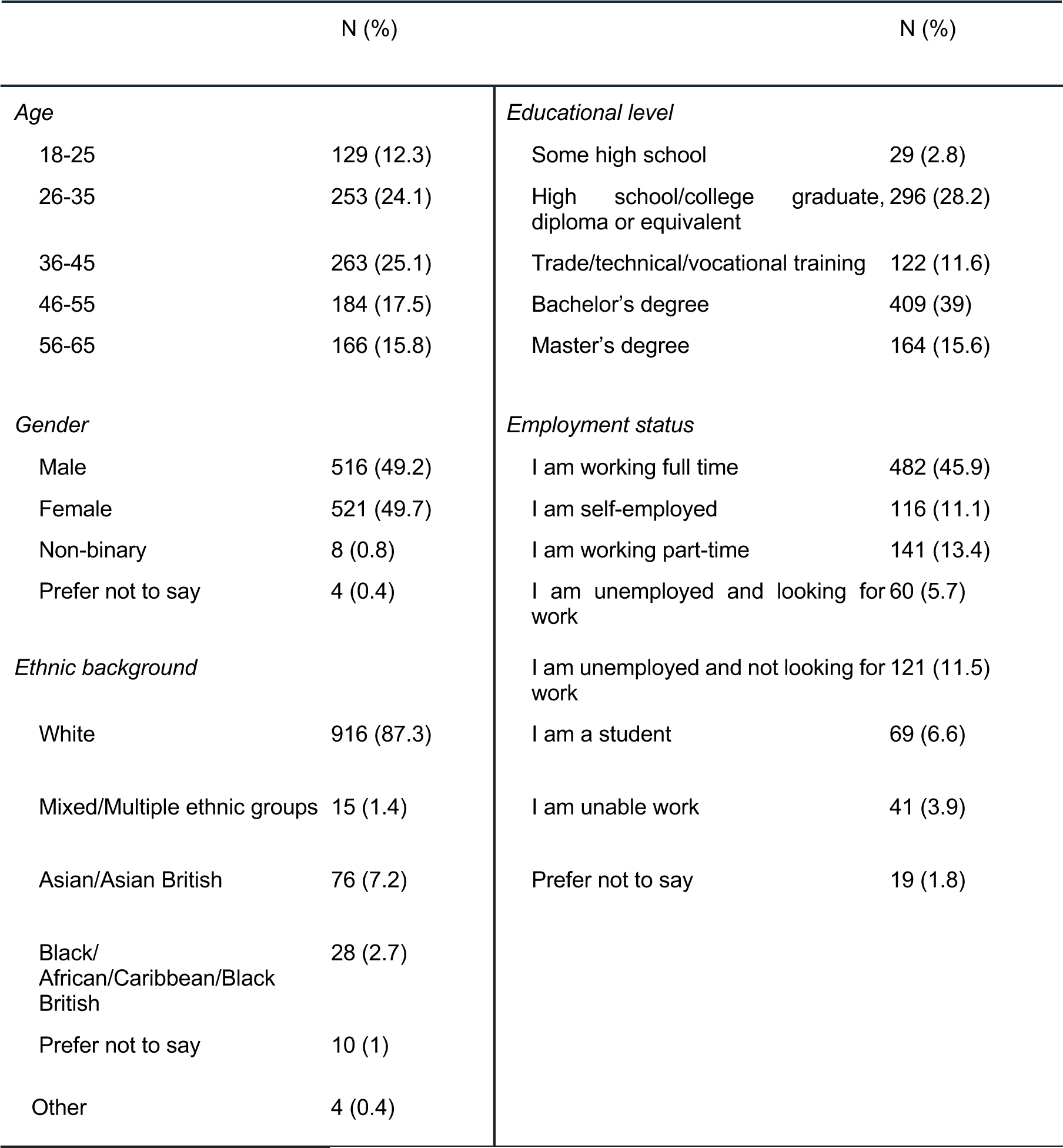
General Population Survey Responder’s Demographic Background.

##### Results

Overall, 91.2% of respondents have never taken part in a clinical trial. However, 68.6% would consider taking part in one, irrespective of the phase. After providing a detailed definition of what an early phase trial entails, 50.2% of responders were willing to consider participation. Focusing on Asian/Asian British responders, 92.9% of the sample had not previously taken part in a clinical trial, but they were slightly more willing to take part in one (71.4%), when compared to the general sample. However, their willingness to take part in an early phase clinical trial was slightly lower than the general sample (48.8%). Regarding Black/African/Caribbean/Black British ethnic group, they were the group with the highest previous participation in a clinical trial, with 81.8% never taking part in a clinical trial, and were somewhat more willing to take part in a clinical trial (72.7%), as well as in an early phase (51.5%; see Figure 2).

**Fig. 2.**
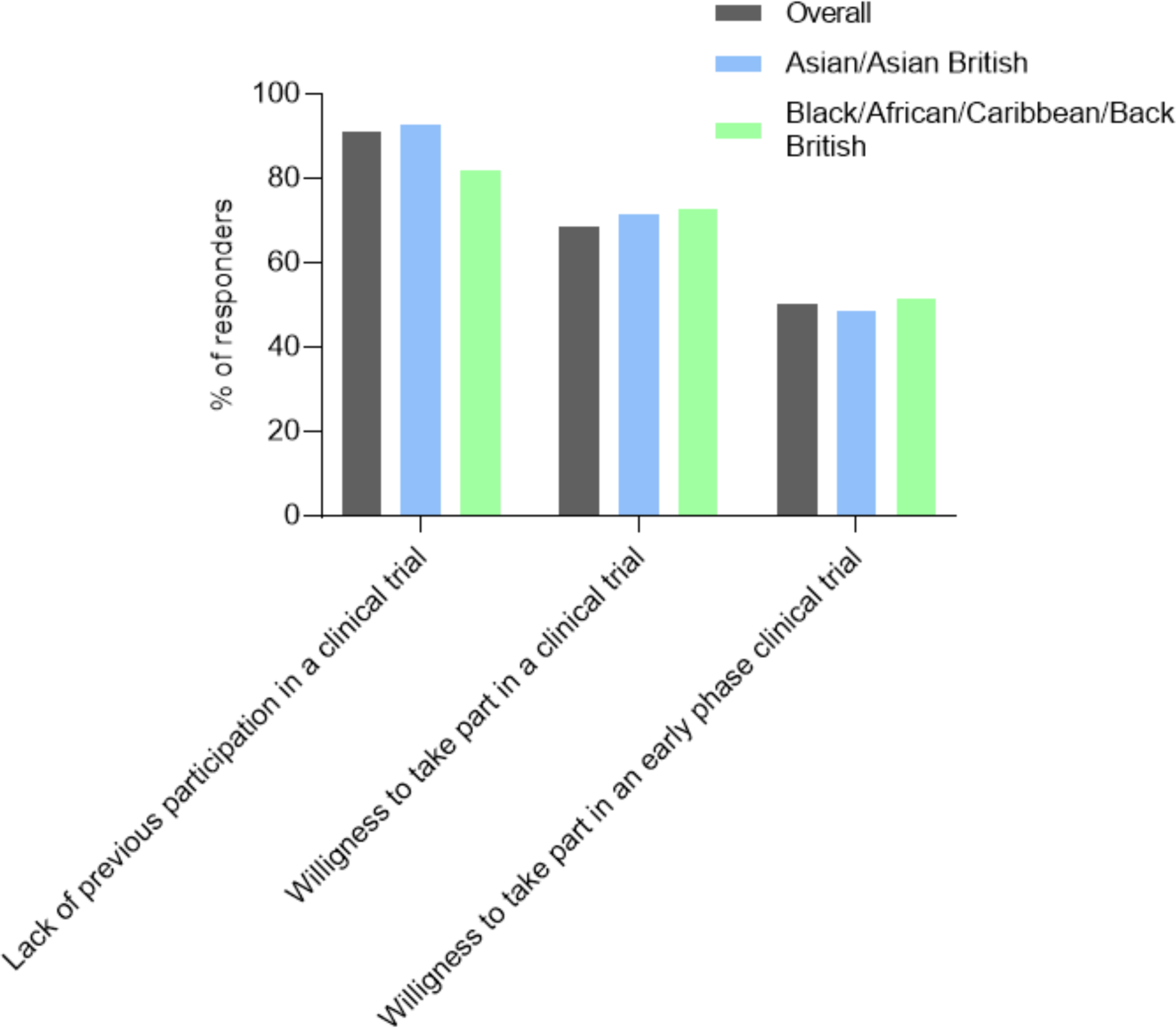
General attitudes and experience towards clinical trials. Previous participation in a clinical trial and their attitudes towards a potential future participation based on participant’s ethnic background.

Overall, the sample reported (i) *financial payment*, (ii) *helping discover new treatments* and (iii) *awareness of the importance of these studies for society* as the key incentives to take part in clinical trials. Interestingly, in early phase clinical trials, participants included *helping discover new treatments* as their top three incentives to take part. This was however different for Asian/Asian British responders, who reported *financial payment* as their top incentive for early phase clinical trial participation. Although Black/African/Caribbean/Black British responders’ incentives matched the general sample, they also included g*eneral interest in science* as an incentive for both early and late phase clinical trials.

Participants selected their top three incentives that motivate and barriers that preclude their participation in a clinical trial, as well as in an early phase clinical trial (see Table 3).

**Table 3.** General Population Survey responders’ top three incentives and concerns for participating in clinical trials (and early phase clinical trials).

| **Ethnic Background** | **Incentives for clinical trials** | **Incentives for early phase clinical trials** | **Concerns for clinical trials** | **Concerns for early phase clinical trials** |
| --- | --- | --- | --- | --- |
| Overall Sample | (1) Financial payment<br>(2) Discover new treatments<br>(3) Awareness of the importance | (1) Discover new treatments<br>(2) Financial payment<br>(3) Awareness of the importance | (1) Safety and health concerns<br>(2) Too much time and effort required<br>(3) Not advised by people around me | (1) Safety and health concerns<br>(2) Not enough financial reimbursement<br>(3) Too much time and effort |
| Asian<br>Asian British | (1) Financial payment<br>(2) Discover new treatments<br>(3) Awareness of the importance for society | (1) Financial payment<br>(2) Discover new treatments<br>(3) Awareness of the importance for society | (1) Safety and Health concerns<br>(2) Too much time and effort<br>(3) Not enough financial payment/No job flexibility | (1) Safety and health concerns<br>(2) Too much time and effort/Not enough financial payment<br>(3) Not advised by people around |
| Black<br>African<br>Caribbean<br>Black British | (1) Financial payment<br>(2) Discover new treatments<br>(3) Interest in Science | (1) Financial payment/Discover new treatments<br>(2) Interest in Science<br>(3) Awareness of importance | (1) Safety and Health concerns<br>(2) Not advised by people around me<br>(3) Not enough financial payment | (1) Safety and health concerns<br>(2) Not enough financial payment<br>(3) Lack of trust |

Regarding common barriers precluding participants from partaking in clinical trials, the sample reported (i) *concerns over safety and health outcomes,* (ii) *amount of time and effort required to take part in one*, and (iii) *not being advised by people around them*. Although somewhat similar for early phase clinical trials, responders reported l*ow monetary reimbursement*, instead of *not being advised by people around them* to be amongst the top three barriers. With Asian/Asian British responders reporting similar concerns as the general population, Black/African/Caribbean/Black British participants reported *lack of trust* as one of their main concerns for early phase clinical trials.

#### Focus groups

##### Participants

A total of 17 participants took part across five focus groups. Participants’ demographic information can be found in Table 4.

**Table 4.** Focus group participants’ demographic background.

|  | N (%) |  | N (%) |
| --- | --- | --- | --- |
| *Age* |  | *Educational level* |  |
| 26-35 | 10 (58.82) | Trade/technical/vocational training | 1 (5.88) |
| 36-45 | 6 (35.29) | Bachelor's degree | 8 (40.06) |
| 56-65 | 1 (5.88) | Master's degree | 7 (41.18) |
| *Gender* |  | Doctorate degree | 1 (5.88) |
| Male | 11 (64.71) | *Ethnic background* |  |
| Female | 6 (35.29) | White | 2 (11.77) |
| *Employment status* |  | Asian/Asian British | 9 (52.94) |
| Working full time | 14 (82.35) | Black/ African/Caribbean/Black British | 6 (35.30) |
| Self-employed | 1 (5.88) |  |  |
| Student | 1 (5.88) |  |  |

In this section we will present the findings from the five focus groups. A total of eight themes emerged from the data: (i) *relevance of the trial*; (ii) *safety confirmation*; (iii) *adequate support/perception from others*; (iv) *monetary compensation*; (v) *trust in the trial sponsor;* (vi) *convenience*; (vii) *awareness of clinical trials and their outcomes*; and (viii) *source of referral*. These themes provide strategies by which to increase uptake among minority ethnic populations for both early and late clinical trials. Given the general overlap between these phases, it is explicitly pointed where these differ between phases.

##### Relevance of the trial

Overall, the relevance that the trial had to participants was an important driver to motivate their participation in a clinical trial. This relevance could apply to (a) themselves or someone close, (b) to their community, with or (c) the wider society.

**Relevance to themselves or someone close:** participants explained that one of the key reasons to take part in a clinical trial is the relevance the trial has to them (Quote a, Supplementary Table 1), and are more willing to take part in a clinical trial if it is for a condition they or someone else close to them is suffering from (Quote b, Supplementary Table 1).

##### Relevance to their community

It was pointed out that clinical trials become more relevant to participants from diverse backgrounds when they aim at ensuring that drugs are tested on a diverse sample, so that it can help people from their community (Quote c, Supplementary Table 1).

##### Relevance to the wider society

Finally, there is a component of relevance when participants are aware that they are contributing to the development of something that would benefit the wider society (Quote d, Supplementary Table 1).

##### Safety confirmation

One aspect that received much attention throughout the discussions was concerns over one’s safety throughout the trial and once the trial is completed (Quote e, Supplementary Table 1). These concerns were not only raised in relation to one’s safety, but also in relation to potential knock-on effects felt by individuals they are responsible for (i.e., kids), or on their jobs (Quote f, Supplementary Table 1).

In early phase clinical trials, these concerns were stronger and harder to overcome, making participants more hesitant to take part. Interestingly, participants expressed that having participated in a late phase clinical trial would increase the chances of them considering taking part in an early phase (Quote g, Supplementary Table 1).

To help with these concerns, participants express that a clear, honest and open communication about what is being tested, its potential impact on the body and side-effects would be reassuring and help with their decision-making process before taking part in a clinical trial (Quote h, Supplementary Table 1). For early phase clinical trials, clarity and transparency of the risk participants will be putting themselves through is very important to their safety throughout the trial, ultimately aiding the decision-making process (Quote i, Supplementary Table 1).

Additionally, adequate care and safety monitoring throughout the trial is also a key aspect for safety reassurance. Participants appreciate having someone with whom they can discuss directly (i.e., a phone number) what they are committing to and contact in case of an adverse event. Similarly, participants appreciate the reassurance that they will receive appropriate care in the case of an adverse event (Quote j, Supplementary Table 1).

##### Monetary compensation

For many participants, money is a big incentive to take part in clinical trials. Participants believed that they should be compensated for travel, time (including time off work), expenses and effort to take part, where effort relates to what is being tested (i.e., device, pill, vaccine, etc.) and how invasive it is for one’s health (risk-related). Congruently, participants expected that early phase clinical trials’ compensation to be higher due to the lack of risk-related information (Quote k, Supplementary Table 1). Participants also expressed the need of additional compensation in case of a serious adverse event (i.e., stroke) or death. Insurance-related concerns were additionally raised (Quote l, Supplementary Table 1).

##### Adequate support/perception from others

Participants consider not only themselves when thinking about taking part in a clinical trial, but also their support networks and other members of their community. Therefore, an adequate perception from others about participating in clinical trials can be considered key in the decision-making process, with negative perceptions held by others acting as a barrier to participation (Quote m, Supplementary Table 1). Such negative perceptions can lead to fear of judgement and rejection from the potential participant, or not being allowed to do so by others (Quote n, Supplementary Table 1). Consistently, making the participant’s support network informed about the research and part of the decision-making process was a strategy suggested to make participants feel safer and as a way to overcome some of the negative but influential perceptions held by other members of their community (Quote o, Supplementary Table 1. Additionally, participants mentioned that if key figures in their community recommended to participate in clinical trials, and were open minded about it, it would be easier to engage more people from that specific community (Quote p, Supplementary Table 1)

##### Trust in the trial sponsor

Participants expressed that their willingness to take part in a clinical trial would depend on the entity that is sponsoring it. Some participants noted that clinical trials sponsored by academic institutions (vs. commercial sponsors) may be more trustworthy. These, as opposed to pharmaceutical companies, were viewed as more credible institutions due to their internal procedures to ensure ethical standards (Quote q, Supplementary Table 1). It was highlighted that whether it was coming from a pharmaceutical company or a university, participants would rather see that the entity running the trial has the experience and knowledge to do so (Quote r, Supplementary Table 1).

##### Convenience

Participants reported that to be able to take part in a clinical trial, it has to be convenient and flexible to be able to accommodate their schedules, which might include 9-5 jobs, responsibilities such as kids, or other commitments (Quote s, Supplementary Table 1). One important aspect of convenience is location, with proximity and accessibility to the trial site being important drivers. Participants expect that if there is some sort of transportation involved in accessing the trial site, this would be paid for by the trial sponsor (Quote t, Supplementary Table 1). Trials with fewer on-site visits were perceived as more convenient, as they could easily accommodate their working patterns. With this, a debate on digital trials opened up. Digital trials were seen as more convenient by some. However, a couple of participants did mention that with digital trials, they would want reassurance of a lower possibility of having side effects (Quote u, Supplementary Table 1).

##### Awareness of trials and their outcomes

There was a general sense of lack of awareness and knowledge about clinical trials until the COVID-19 pandemic. Additionally, what participants reported to know about them was marked by a negative perception, mainly arising from the negative media that had spread after specific adverse events happening in clinical trials (Quote v, Supplementary Table 1). Participants reported that these negative perceptions could be tackled by spreading pieces of media highlighting what has been achieved through clinical trials (Quote w, Supplementary Table 1). The lack of information available to participants about clinical trials is therefore impacting current recruitment practices. Consequently, when potential participants are approached through media advertisements about a clinical trial, their lack of awareness about them together with ads not providing sufficient information, makes it hard for participants to make an informed decision (Quote x, Supplementary Table 1).

To target different parts of the population, participants agree that spreading awareness and advertising the studies in community centres, such as an influential individual within the community or using their news channels, would be a great space to advertise and let individuals know about the existence of these (Quote y, Supplementary Table 1). Finally, participants mentioned that they would highly appreciate hearing the feedback of what they were able to contribute to (Quote z, Supplementary Table 1).

##### Source of referral

Trust in who is referring the clinical trial was an important factor when participants were considering their participation, with clinical trials referred by GPs or medical doctors perceived as highly trusted (Quote aa and ab, Supplementary Table 1). Participants also remarked that hearing the experience of taking part in a clinical trial from someone that has already done it would ease the decision-making process. This ranged from previous participants within their community recommending the trial, to having the opportunity to meet and discuss the trial and the experience of taking part in it with someone that has already done it (Quote ac, Supplementary Table 1).

Altogether, these seven themes report those aspects that incentivise participation and those that act as barriers. In case of the latter, strategies derived from the data were proposed to overcome these.

## Discussion

This study used a mixed methodological approach to explore barriers that discourage ethnically diverse individuals from participating in clinical trials, with a specific focus on early phase clinical trials. Findings showed that (i) little research has been conducted within the domain (Systematic Review), (ii) there is a discordance between what researchers and the general population perceive as barriers to participation (e-surveys), and (iii) eight factors or strategies may encourage participation in ethnically diverse populations (Focus Groups). Considered holistically, findings from this investigation indicate a *Clinical Trials Participatory Framework* that contextualises reduced participation, while providing a model to increase uptake among minority ethnic populations.

The systematic review revealed there was very little evidence-based research that considered barriers to diverse and representative participation in early phase clinical trials. Given this knowledge-based gap, it was considered timely to gauge researcher’s and an ethnically diverse population’s perceptions of factors that may inhibit participation in clinical studies. Findings showed discordance between these groups with regard to both drivers and barriers to participation. While researchers cited practical and procedural related difficulties (i.e., *complicated study requirements*, *non-compatible schedules*, d*ifficulties with consent*), the general population survey indicated *safety and health concerns* as impacting research involvement, particularly in early phase trials. This observation is consistent with previous work showing that *safety and health concerns*, which are exacerbated and highly influenced by media coverage, shape participant attitudes and create reticence, especially in early-stage research (20). Participants indicated that uncertainties about risk and safety were too often minimised and a clearer and transparent discussion of these issues would build trust. Where risks had been identified, participants wanted to hear about mitigation strategies and be reassured that appropriate medical care would be available if problems arose.

Both surveys showed a further mismatch as to the relevance and importance of *financial reimbursement* to trial participation. Participants were clear that they considered that the perceived greater risk of early stage research should attract enhanced monetary reimbursement, while *poor or lack of monetary reward* constituted the least important barrier identified by researchers. The issue of financial reimbursement has been a subject of much debate within the medical community. Those leading and regulating research have been clear that financial reward should only be reflective of time and inconvenience. Risk-related payment is considered inappropriate as such payments may be construed as paternalistic and potentially diminish a participant’s ability to make an informed decision (21) On the other hand, physicians have recognised that payment should include an aspect of risk, to compensate and mitigate against risk-related outcomes, particularly in early phase research (22) Despite participants’ *interest in science* and *awareness of research importance* somewhat mitigating issues related to limited financial reimbursement, the persistent mismatch between participants and researchers’ views represents an endemic problem with existing recruitment practices. Given this gap both in understanding and practice, the focus groups explored strategies to incentivise participation amongst minority ethnic groups and provide researchers with a framework to improve recruitment processes.

Based upon the drivers and barriers to participation identified by minority ethnic individuals, findings from the focus groups provide initial evidence for the support of a *Clinical Trials Participatory Framework* (‘framework’). The framework seeks to both contextualise reasons for reduced trial participation, while providing mechanisms and strategies by which to increase uptake among minority ethnic populations. Specifically, the framework identifies ‘eight barriers turned incentives’ regarded by participants as integral to improving trial participation among diverse populations. As shown in Figure 3, the framework progresses as a 2-phased pathway, with each phase acting as a potential gatekeeper to trial participation, while feedback loops continuously act to either affirm or contradict a participant’s decision. Within Phase 1 of this framework, *relevance of the trial, monetary compensation* and *convenience* can be seen as foundational factors. These factors act to shape participant’s initial perceptions regarding the importance and feasibility of their participation within the study (i.e., “Is my participation cost-effective”, “Do I or someone I know gain something from my participation?”). The existence of one or more of these factors is necessary for an individual to progress to Phase 2. Once an initial participatory drive or willingness has been established (Phase 1), individuals undertake a second implicit assessment of ‘trustworthiness’ (Phase 2), which considers the *reputation and trust of the trial sponsor, source of referral*, *support/perception from others,* and their own understanding and *awareness of the trial topic/area*. This trustworthiness assessment safeguards against taking part in dubious, questionable, or unethical trials (i.e., unknown spam source) that may oppose their values or lead to judgement by others. Irrespective of the phase, individuals confirmed undertaking background *safety confirmations* to insulate against adverse physical or psychological events, with these ‘checks’ acting as continuous feedback loops to either affirm or contradict a particular decision. More simply, if the clinical trial satisfies one or more of the foundation factors, and is deemed both safe and trustworthy, then the individual will likely take part in the research. However, if the study is deemed important but either unsafe or untrustworthy, the individual will likely not take part in the research, unless one of the underlying reasons identified as part of Phase 1 (i.e., monetary reimbursement, relevance of trial) outweigh the perceived risks.

**Fig 3.**
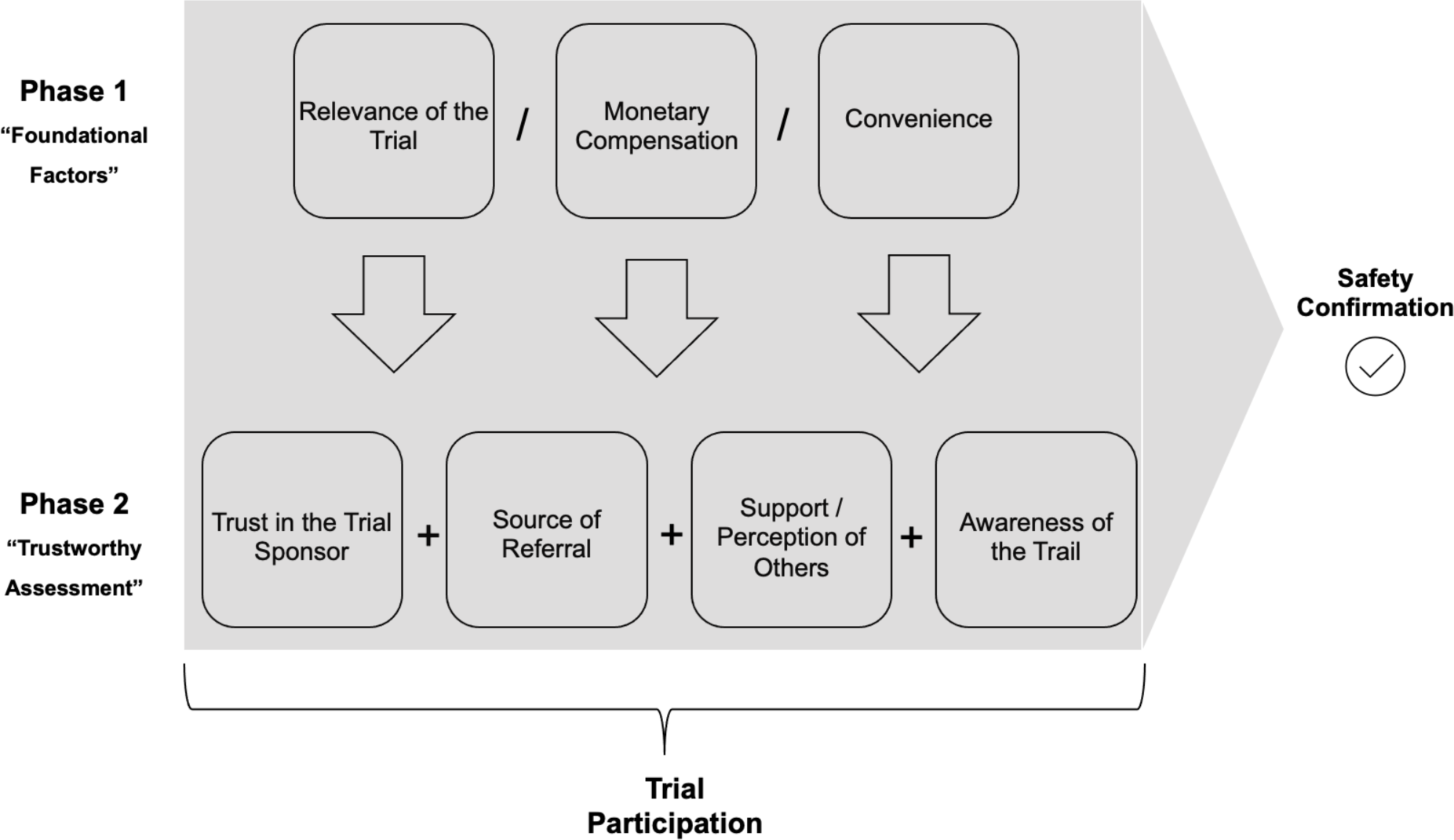
Clinical Trials Participatory Framework. A 2-phased pathway, with each phase acting as a potential gatekeeper to trial participation. Starting from Phase 1, “Foundational factors” shape participant’s initial perceptions regarding the importance and feasibility of their participation in the trial. The existence of one or more of these factors is necessary for an individual to progress to Phase 2. Once Phase 1 is established, potential participants undertake a “Trustworthy Assessment” as part of Phase 2 that safeguards them taking part in dubious, questionable, or unethical trials that may oppose their values or lead to judgement by others. Irrespective of the phase, *safety confirmation* checks continuously act as feedback loops to either affirm or contradict a particular decision.

The *Clinical Trials Participatory Framework* provides a model through which to understand the decision-making processes employed by potential participants, while offering researchers eight key strategies to improve recruitment and participation in early and late- phase clinical research.

### Strengths, limitations and future directions

The tripartite nature of the investigation (i.e., systematic review, e-surveys, focus groups), provided rich qualitative data, allowing for in-depth exploration. We consider that aligning researcher and participants perspectives is vital to enable wider participation, and that the *Clinical Trials Participatory Framework* provides a practical means to structure more effective recruitment strategies.

Limitations include limited sample sizes (i.e., researcher survey), together with sampling variability across groups, which may limit the generalisability of the stated results. Nevertheless, these findings provide a practical and strategic approach to guide researchers working within this domain. Future research should evaluate the effectiveness of the proposed framework with the aim of enabling iterative refinement.

## Conclusion

To the best of the researcheŕs knowledge, this study constitutes one of the first explorations of barriers that discourage ethnically diverse individuals from participating in early phase clinical trials. Findings from this tripartite investigation provided the basis for a *Clinical Trials Participatory Framework* which contextualises reduced participation while encouraging uptake among ethnically diverse populations. Through the use of this framework, we hope that parity of participation amongst ethnically diverse groups and the general population may be reached.

## Declarations

### Ethics approval and consent to participate

It was determined by the project steering committee that an ethics committee application was not needed for the online surveys nor focus groups as these falls under the category of “market research”. This was informed using the HRA Research Decision Tool (http://www.hra-decisiontools.org.uk/research/).

### Consent for publication

Not applicable

### Availability of data and materials

The datasets used and/or analysed during the current study are available from the corresponding author on reasonable request. However, focus groups recordings obtained during the current study will not be publicly available in order to preserve anonymity.

### Authors’ contributions

PAH prepared the surveys, focus group content, acquired and analysed the data, drafted the manuscript and prepared it for submission. NM conducted and drafted the systematic review. PW helped plan the project and contributed to the development of the surveys and focus group content, provided support for the analysis of the data. RB contributed to the development of the surveys and focus group content, recruitment. MY helped plan the project and contributed to the development of the surveys and focus group content. DR acquired focus group data. All authors critically revised the manuscript and approved for submission.

## Data Availability

All data produced in the present work are contained in the manuscript

## Acknowledgements

We want to acknowledge the EDICT advisory committee, who helped plan the project and review the content of the surveys, focus groups, and manuscript. This committee is composed of Dr Ly-Mee Yu, Professor Paramjit Gill, Dr Lydia Poole, Dr Mel Ramasawmy and Dr Mariam Molokhia.

